# A Novel Risk Assessment Model Predicts Major Bleeding Risk at Admission in Medical Inpatients

**DOI:** 10.1101/2023.04.29.23289304

**Authors:** Benjamin G Mittman, Megan Sheehan, Lisa Kojima, Nicholas Cassachia, Oleg Lisheba, Bo Hu, Matthew Pappas, Michael B. Rothberg

**Affiliations:** Center for Value-Based Care Research, Cleveland Clinic, Cleveland, OH; Department of Population and Quantitative Health Sciences, Case Western Reserve University, Cleveland, OH; Enterprise Analytics eResearch Department, Cleveland Clinic, Cleveland, OH; Department of Quantitative Health Sciences, Cleveland Clinic, Cleveland, OH

**Keywords:** bleeding, risk assessment model, clinical prediction, machine learning

## Abstract

**Background:** Venous thromboembolism (VTE) is the leading cause of preventable hospital death in the US. Guidelines from the American College of Chest Physicians and American Society for Hematology recommend providing pharmacological VTE prophylaxis to acutely or critically ill medical patients at acceptable bleeding risk, but there is currently only one validated risk assessment model (RAM) for estimating bleeding risk. We developed a RAM using risk factors at admission and compared it with the International Medical Prevention Registry on Venous Thromboembolism (IMPROVE) model.

**Methods:** A total of 46,314 medical patients admitted to a Cleveland Clinic Health System hospital from 2017-2020 were included. Data were split into training (70%) and validation (30%) sets with equivalent bleeding event rates in each set. Potential risk factors for major bleeding were identified from the IMPROVE model and literature review. Penalized logistic regression using LASSO was performed on the training set to select and regularize important risk factors for the final model. The validation set was used to assess model calibration and discrimination and compare performance with IMPROVE. Bleeding events and risk factors were confirmed through chart review.

**Results:** The incidence of major in-hospital bleeding was 0.58%. Active peptic ulcer (OR = 5.90), prior bleeding (OR = 4.24), and history of sepsis (OR = 3.29) were the strongest independent risk factors. Other risk factors included age, male sex, decreased platelet count, increased INR, increased PTT, decreased GFR, ICU admission, CVC or PICC placement, active cancer, coagulopathy, and in-hospital antiplatelet drug, steroid, or SSRI use. In the validation set, the Cleveland Clinic Bleeding Model (CCBM) had better discrimination than IMPROVE (0.86 vs. 0.72, p < .001) and, at equivalent sensitivity (54%), categorized fewer patients as high-risk (6.8% vs. 12.1%, p < .001).

**Conclusions:** From a large population of medical inpatients, we developed and validated a RAM to accurately predict bleeding risk at admission. The CCBM may be used in conjunction with VTE risk calculators to decide between mechanical and pharmacological prophylaxis for at-risk patients.

## Introduction

Venous thromboembolism (VTE) is the leading cause of preventable hospital death in the United States (US).^1^ Multiple randomized controlled trials have demonstrated that pharmacological prophylaxis with heparin is highly effective at reducing the incidence of VTE and subsequent mortality.^2,3^ However, heparin also increases the risk of bleeding.^3^ Therefore, the American College of Chest Physicians (ACCP)^4^ and American Society for Hematology (ASH)^5^ recommend that medical patients be assessed for risk of VTE and those not at low risk receive pharmacological VTE prophylaxis, but only if they are at acceptable bleeding risk.

However, assessment of bleeding risk can be challenging^6^ and the guidelines do not specify how bleeding risk should be determined. Unlike assessment of VTE, for which there are multiple validated risk assessment models (RAMs),^7–11^ there is only one validated RAM available for estimating bleeding risk, the International Medical Prevention Registry on Venous Thromboembolism (IMPROVE) model.^12^ Originally derived on 9,388 patients from hospitals in 12 countries using multiple logistic regression, IMPROVE includes 11 factors at admission that were independently associated with in-hospital major or non-major but clinically relevant bleeding. A risk score was developed to assign points for each factor, allowing categorization of patients as either high- or low-risk for bleeding based on their total score.

Despite acceptable performance in the original data set, IMPROVE has key limitations. These include insufficient validation of risk factor data extracted from the electronic health records (EHRs), informal variable selection, exclusion of several important potential risk factors, and inclusion in the outcome of non-major bleeds, which although clinically relevant, typically do not warrant modification of VTE prophylaxis. Additionally, although IMPROVE has been externally validated, its performance in validation studies has been poor.^13,14^ In the largest validation study, approximately 20% of patients were categorized as high risk, sensitivity was less than 40%, and the area under the receiver operating characteristics curve (AUROC) was 0.63.^13^

A more accurate and generalizable bleeding RAM is needed to better evaluate bleeding risk at admission in medical inpatients. We identified major in-hospital bleeds and derived a RAM based on more than 30,000 patients from 10 hospitals within the Cleveland Clinic Health System (CCHS). We then validated our RAM on a holdout validation set of nearly 14,000 patients and compared its discrimination and calibration with the IMRPOVE model.

## Methods

### Setting and Patients

Our cohort consisted of medical inpatients who were admitted to one of 10 CCHS hospitals between October 1, 2017, and January 31, 2020. Hospitals were located in Ohio and Florida and varied in size from a 126-bed community hospital to a 1,400-bed quaternary care academic medical center. We included patients aged 18 years or older who were admitted to a medical service, either directly or from the emergency room (ER). Because we were interested in hospital-acquired bleeding, we excluded patients who had a bleed that was either documented as present-on-admission or occurred within 48 hours of admission. We also excluded patients taking warfarin because they would not be eligible to receive chemoprophylaxis. The study was approved by the Cleveland Clinic Institutional Review Board (IRB #14-240).

### Identification of Bleeds

We developed an algorithm to identify major in-hospital bleeds in medical inpatients. We used the International Society on Thrombosis and Hemostasis (ISTH) definition of major bleeding—clinically overt and either fatal or associated with one of the following: (a) fall in hemoglobin of 2 g/dL or more, (b) documented transfusion of at least 2 units of packed red blood cells, or (c) involvement of a critical anatomical site (e.g., intracranial, pericardial, intramuscular with compartment syndrome, retroperitoneal).^15^ Additionally, the bleed could not be perioperative and must have resulted in expected blood loss. We used a combination of diagnostic codes and laboratory values to identify patients with probable bleeds based on these criteria. We reviewed the charts of patients with probable bleeds to confirm all outcomes. We excluded patients with a present-on-admission indicator for a bleeding code, bleed during admission, or major surgical operation without a specific diagnostic code for other bleeding.

### Risk Factor Identification

We identified potential risk factors for major bleeding from the IMPROVE^12^ model and other published studies.^16–20^ We considered age, sex, race, smoking status, alcohol use, platelet count, international normalized ratio (INR), partial thromboplastin time (PTT), glomerular filtration rate (GFR), prior bleeding within 3 months of admission, rheumatic disease, sepsis, intensive care unit (ICU) admission within 24 hours of hospital admission, central venous catheter (CVC) placement (including peripherally inserted central catheter, PICC) within 24 hours of admission, cancer, peptic ulcer, in-hospital antiplatelet medication use, hemorrhagic or ischemic stroke, transient ischemic attack, hypertension, heart failure, pericarditis, diabetic retinopathy, hospital readmission, and myocardial infarction. All variables were extracted from the Cleveland Clinic EHR system.

Smoking status and alcohol use were categorized as current, former, or never. Platelet count (thousands of platelets per uL), INR, and PTT (seconds) values were not modified. Serum creatinine (umol/L) was used to calculate GFR (mL/min) using the revised chronic kidney disease-epidemiology age and sex (CKD-EPIAS) equation.^21^ Antiplatelet medications (aspirin, clopidogrel, ticagrelor, or prasrugrel) were categorized as yes/no. Readmission was defined as having a previous inpatient discharge within 30 days of admission. All other predictors were identified based on a combination of diagnostic and finance codes (see Supplementary Information).

Because many patients received chemoprophylaxis, which is hypothesized to increase the risk of major bleeding,^17^ we identified VTE prophylaxis (subcutaneous heparin 5,000 units 2–3 times daily, enoxaparin 40mg daily, dalteparin 5,000 units daily, or fondaparinux 2.5mg daily) from the table of medications administered. To assess the impact of prophylaxis on model parameter estimates, we performed a sensitivity analysis, refitting a multiple logistic regression model with and without patients who received prophylaxis.

Data extracted from the EHR are often unreliable,^22^ thus we performed iterative chart review of a random sample of patient charts for each predictor. To ensure that each risk factor preceded the outcome, we required that all predictors be present in the first 24 hours of admission.

### Model Development and Training

We first performed single imputation to fill in missing values for INR and PTT based on the distribution of values in patients with normal results. We did not perform multiple imputation because these data were not missing at random. Unlike platelet count and serum creatinine (used to calculate GFR), INR and PTT are not routinely measured in hospitalized patients unless there are specific indicating symptoms or suspected bleeding or coagulation disorders. Thus, we assumed that patients with missing INR and PTT values had values within normal range.

We split the cohort into training (70%) and validation (30%) sets with pseudo-randomization of bleeding events to ensure proportional event rates in each set. We did this to ensure model trainability and accuracy in the presence of a highly unbalanced binary outcome. We used the training set to perform variable selection in two steps. First, we used univariate analyses (t-tests and chi-squared tests for continuous and categorical variables, respectively) and collinearity assessments among the full set of predictors to exclude unimportant variables. We then used a supervised machine learning algorithm, the least absolute shrinkage selection operator (LASSO),^23^ to identify remaining unimportant variables in order to minimize model complexity and reduce overfitting. Variables with no importance in predicting major bleeding were penalized and shrunk to zero and removed from the final fitted model. Included variables were regularized based on their amount of shrinkage. Cross-validation was used to determine the penalty term, lambda, and to compute the regularized coefficient for each variable. Coefficients were exponentiated to yield odds ratios (ORs).

All analyses were performed in R (v4.2.2).^24^ LASSO regression modeling was performed using the package glmnet (v4.1-6).^25^ In R, glmnet automatically centers and scales all data prior to fitting the LASSO model, thus we did not standardize any variables. However, we scaled down age and platelet count by a factor of 10 to reduce variability in the range of scales between predictors (since most were binary and coded as 0 or 1) and to facilitate easier interpretation of odds ratios.

### Internal Validation

The LASSO model fit on the training set was used to generate a predicted probability of bleeding for each patient in the validation set. Model calibration was assessed using a calibration plot. Model discrimination was assessed using a receiver operating characteristics (ROC) curve, the area under the ROC curve (AUROC), sensitivity, specificity, and the F1 score (harmonic mean). The optimal cut-off for predicting bleeding was chosen as the point on the final ROC curve that maximized the Youden index. Bootstrapping was used to compute the optimism-adjusted AUC and 95% confidence interval (CI) at the optimal cut-off, as well as to plot 95% confidence bands for the ROC curve at a range of sensitivity and corresponding specificity values.

### External Validation and Comparison with IMPROVE

We sought to validate the IMPROVE model in an external sample and compare the performance of our LASSO model (hereafter Cleveland Clinic Bleeding Model [CCBM]) with the IMPROVE model. To do this, we fit the IMPROVE model on our validation set using multiple logistic regression^12^ and generated predictions in the validation set. We then compared CCBM with IMPROVE on the basis of calibration and discrimination in the validation set. Specifically, we compared models on bleed detection and high-vs. low-risk categorization rates using chi-squared tests, and on AUC using DeLong’s test for paired ROC curves.^26^

In order to compare expected performance of the models in clinical practice, we determined the cut-off on the CCBM that corresponds to a total risk score of ≥7 on the IMPROVE model. We then compared F1 scores as well as high-vs. low-risk categorization rates between the models using a chi-square test. Statistical significance was defined as two-sided p < .05.

### Comparing CCBM with Clinician Assessment

The purpose of developing this RAM was to design a tool that could aid clinician decision-making for VTE and bleeding risk in hospitalized patients. We empirically assessed clinicians’ decisions for VTE prophylaxis use in patients stratified by bleeding risk to determine how well clinicians incorporate bleeding risk assessment into prophylaxis decision-making at baseline. We divided patients into deciles based on bleeding risk computed by CCBM and quantified the percentage of eligible patients who received prophylaxis in each decile. Patients ineligible for chemoprophylaxis were those whom clinicians considered to be at high risk of bleeding and were excluded from this analysis.

## Results

We identified 48,030 adult medical inpatients with valid bleeding data during the study period. After excluding patients with one or more missing predictors that could not be imputed because the reason for missing values could not be determined, our final cohort contained 46,314 patients. The training set included 32,419 patients (70%) and the validation set included 13,895 patients (30%); each had a major bleed rate of 0.58%. Compared with patients without major bleeding, patients with major bleeds were older (mean age 64.5 vs. 61.4 years); had lower platelet counts (mean count 197,000 vs. 231,000 per uL) and worse renal function (mean GFR 61.5 vs. 80.7 ml/min); and were more likely to have prior bleeding (25.7% vs. 5.9%), sepsis (50% vs. 14.2%), and cancer (28% vs. 15.4%). All patient characteristics and their relationship to major bleeding are shown in Table 1.

**Table 1.**
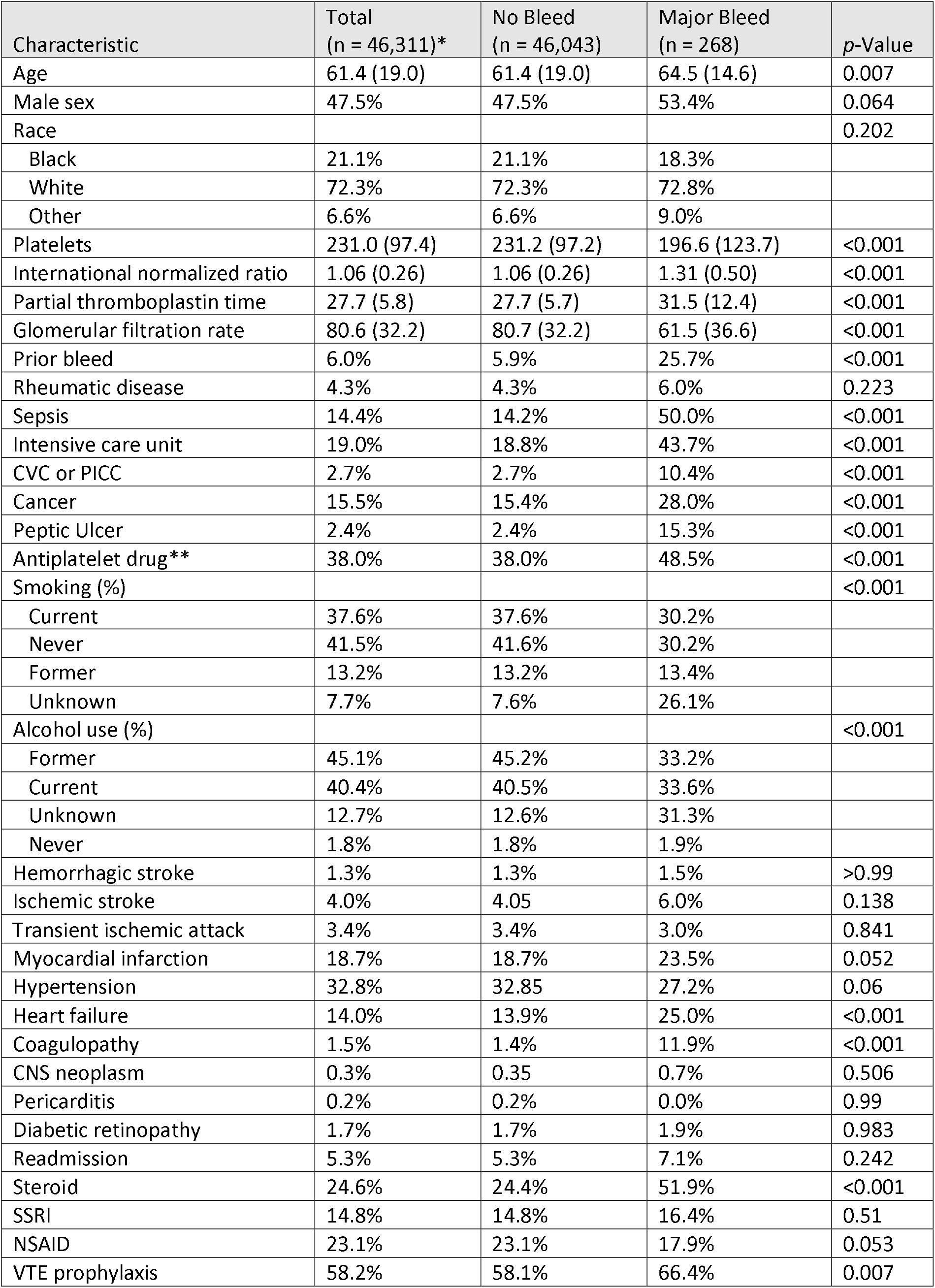
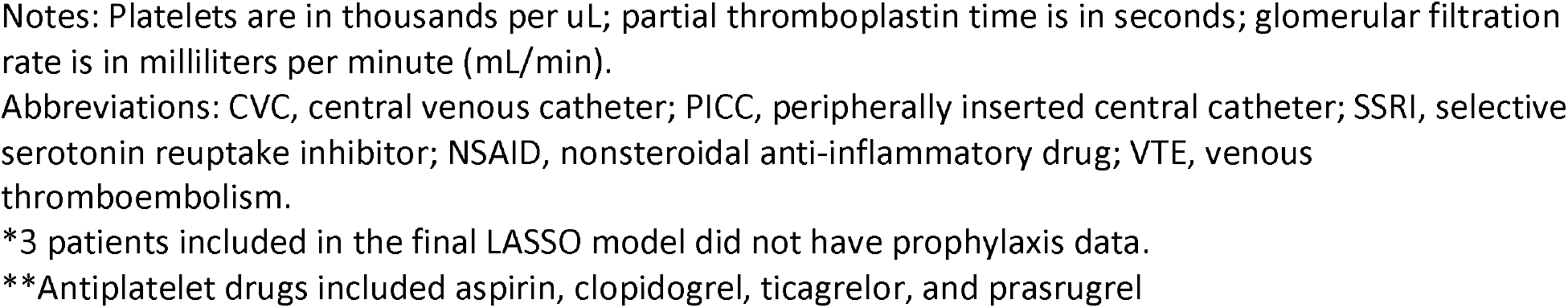
Patient characteristics and their relationship to major in-hospital bleeding.

| Characteristic | Total<br>(n = 46,311)* | No Bleed<br>(n = 46,043) | Major Bleed<br>(n = 268) | p-Value |
| --- | --- | --- | --- | --- |
| Age | 61.4 (19.0) | 61.4 (19.0) | 64.5 (14.6) | 0.007 |
| Male sex | 47.5% | 47.5% | 53.4% | 0.064 |
| Race |  |  |  | 0.202 |
| Black | 21.1% | 21.1% | 18.3% |  |
| White | 72.3% | 72.3% | 72.8% |  |
| Other | 6.6% | 6.6% | 9.0% |  |
| Platelets | 231.0 (97.4) | 231.2 (97.2) | 196.6 (123.7) | <0.001 |
| International normalized ratio | 1.06 (0.26) | 1.06 (0.26) | 1.31 (0.50) | <0.001 |
| Partial thromboplastin time | 27.7 (5.8) | 27.7 (5.7) | 31.5 (12.4) | <0.001 |
| Glomerular filtration rate | 80.6 (32.2) | 80.7 (32.2) | 61.5 (36.6) | <0.001 |
| Prior bleed | 6.0% | 5.9% | 25.7% | <0.001 |
| Rheumatic disease | 4.3% | 4.3% | 6.0% | 0.223 |
| Sepsis | 14.4% | 14.2% | 50.0% | <0.001 |
| Intensive care unit | 19.0% | 18.8% | 43.7% | <0.001 |
| CVC or PICC | 2.7% | 2.7% | 10.4% | <0.001 |
| Cancer | 15.5% | 15.4% | 28.0% | <0.001 |
| Peptic Ulcer | 2.4% | 2.4% | 15.3% | <0.001 |
| Antiplatelet drug** | 38.0% | 38.0% | 48.5% | <0.001 |
| Smoking (%) |  |  |  | <0.001 |
| Current | 37.6% | 37.6% | 30.2% |  |
| Never | 41.5% | 41.6% | 30.2% |  |
| Former | 13.2% | 13.2% | 13.4% |  |
| Unknown | 7.7% | 7.6% | 26.1% |  |
| Alcohol use (%) |  |  |  | <0.001 |
| Former | 45.1% | 45.2% | 33.2% |  |
| Current | 40.4% | 40.5% | 33.6% |  |
| Unknown | 12.7% | 12.6% | 31.3% |  |
| Never | 1.8% | 1.8% | 1.9% |  |
| Hemorrhagic stroke | 1.3% | 1.3% | 1.5% | >0.99 |
| Ischemic stroke | 4.0% | 4.05 | 6.0% | 0.138 |
| Transient ischemic attack | 3.4% | 3.4% | 3.0% | 0.841 |
| Myocardial infarction | 18.7% | 18.7% | 23.5% | 0.052 |
| Hypertension | 32.8% | 32.85 | 27.2% | 0.06 |
| Heart failure | 14.0% | 13.9% | 25.0% | <0.001 |
| Coagulopathy | 1.5% | 1.4% | 11.9% | <0.001 |
| CNS neoplasm | 0.3% | 0.35 | 0.7% | 0.506 |
| Pericarditis | 0.2% | 0.2% | 0.0% | 0.99 |
| Diabetic retinopathy | 1.7% | 1.7% | 1.9% | 0.983 |
| Readmission | 5.3% | 5.3% | 7.1% | 0.242 |
| Steroid | 24.6% | 24.4% | 51.9% | <0.001 |
| SSRI | 14.8% | 14.8% | 16.4% | 0.51 |
| NSAID | 23.1% | 23.1% | 17.9% | 0.053 |
| VTE prophylaxis | 58.2% | 58.1% | 66.4% | 0.007 |

Notes: Platelets are in thousands per $\mu\text{L}$ ; partial thromboplastin time is in seconds; glomerular filtration rate is in milliliters per minute ( $\text{mL/min}$ ).
Abbreviations: CVC, central venous catheter; PICC, peripherally inserted central catheter; SSRI, selective serotonin reuptake inhibitor; NSAID, nonsteroidal anti-inflammatory drug; VTE, venous thromboembolism.
\*3 patients included in the final LASSO model did not have prophylaxis data.
\*\*Antiplatelet drugs included aspirin, clopidogrel, ticagrelor, and prasugrel

The final LASSO model included 16 predictors; see Table 2 for predictors and regularized ORs generated in the training set. The strongest predictors of major bleeding were peptic ulcer (OR = 5.90), prior bleed (OR = 4.24), and sepsis (OR = 3.29). In the validation set, the optimism-adjusted AUC for CCBM was 0.86 (95% CI, 0.81-0.91). At the optimal cut-off, defined as the Youden index (0.68%), sensitivity was 79%, specificity was 84%, and the F1 score was 0.055. Calibration was good (Figure 1). Predicted risk of major bleeding among all patients in the validation set ranged from 0.017% to 58%, with a mean of 0.58% (SD = 1.75%).

**Table 2.** Multivariable and LASSO models for major in-hospital bleeding.

| Characteristic | OR excluding prophylaxis (n = 19,365) | OR including prophylaxis (n = 46,311) | OR including prophylaxis (95% CI) |  | OR including prophylaxis, LASSO (n = 46,314) |
| --- | --- | --- | --- | --- | --- |
| Age* | 0.91 | 0.87 | 0.81 | 0.95 | 0.91 |
| Male Sex | 0.86 | 1.02 | 0.80 | 1.30 | 1.16 |
| Platelets* | 0.97 | 0.98 | 0.97 | 0.99 | 0.98 |
| International normalized ratio | 1.16 | 1.23 | 1.00 | 1.48 | 1.31 |
| Partial thromboplastin time | 1.23 | 1.23 | 1.11 | 1.36 | 1.33 |
| Glomerular filtration rate | 0.87 | 0.89 | 0.85 | 0.92 | 0.91 |
| Prior Bleed | 4.58 | 3.93 | 2.94 | 5.20 | 4.24 |
| Sepsis | 3.53 | 3.36 | 2.60 | 4.33 | 3.29 |
| Intensive care unit | 1.24 | 1.61 | 1.23 | 2.11 | 1.78 |
| CVC or PICC | 1.22 | 1.01 | 0.66 | 1.52 | 0.71 |
| Cancer | 2.03 | 1.60 | 1.21 | 2.11 | 1.83 |
| Peptic Ulcer | 3.06 | 5.81 | 3.99 | 8.26 | 5.90 |
| Antiplatelet | 2.27 | 1.84 | 1.42 | 2.37 | 1.57 |
| Coagulopathy | 3.53 | 2.46 | 1.54 | 3.84 | 1.87 |
| Steroid | 2.94 | 2.69 | 2.11 | 3.43 | 2.61 |
| SSRI | 1.65 | 1.11 | 0.79 | 1.53 | 1.14 |
| NSAID | - | - | - | - | 1.00 |
| VTE prophylaxis | - | 1.37 | 1.05 | 1.80 | - |

Abbreviations: OR, odds ratio; LASSO, least absolute shrinkage selection operator; CI, confidence interval; CVC, central venous catheter; PICC, peripherally inserted central catheter; SSRI, selective serotonin reuptake inhibitor; NSAID, nonsteroidal anti-inflammatory drug; VTE, venous thromboembolism.
\*Scaled down by a factor of 10 (e.g., from years to tens of years)

**Figure 1.**
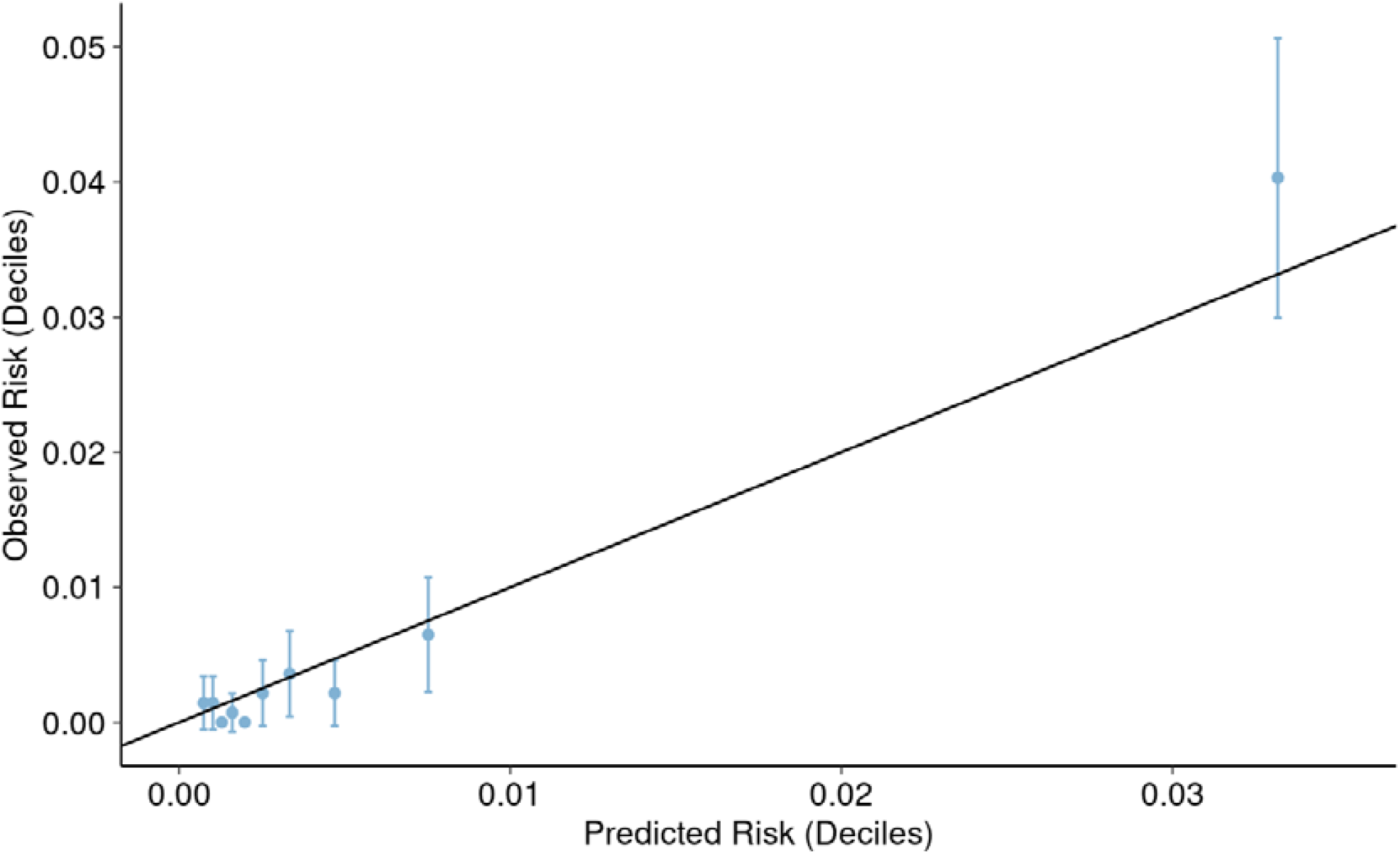
Calibration plot for the Cleveland Clinic Bleeding Model in the validation set.

### Sensitivity Analysis

In addition to fitting the LASSO model, we fit multiple logistic regression models on the entire cohort with and without patients who received VTE prophylaxis to assess the impact of prophylaxis on model parameter estimates. Peptic ulcer’s effect was significantly reduced in the model excluding patients who received prophylaxis (OR = 3.06) compared with the full cohort model (OR = 5.81, 95% CI: 3.99, 8.26), but all other variables did not differ significantly between models (Table 2). Based on these results, we kept all patients with complete data in the final LASSO model, regardless of prophylaxis receipt.

### External Validation of IMRPOVE

The optimism-adjusted AUC for IMPROVE was 0.72 (95% CI, 0.66-0.78). Using the IMPROVE cut-off of ≥7 points, sensitivity was 54%, specificity was 88%, and the F1 score was 0.050. Calibration was generally good but IMPROVE tended to over-predict patients’ risk in the top 3 deciles (Figure S1).

### Comparing CCBM with IMPROVE

The ROC curves with 95% confidence bands are compared in Figure 2. At most values of sensitivity and corresponding specificity, including at the optimal cut-off point for IMPROVE (defined as ≥7 points), CCBM showed better discrimination than IMPROVE. The overall difference was robust; CCBM had a significantly higher area under the curve (AUC) compared with IMPROVE (0.86 vs. 0.72, Z = 4.20, p < .001; 95% CI for the difference between AUCs = 0.08-0.21).

**Figure 2.**
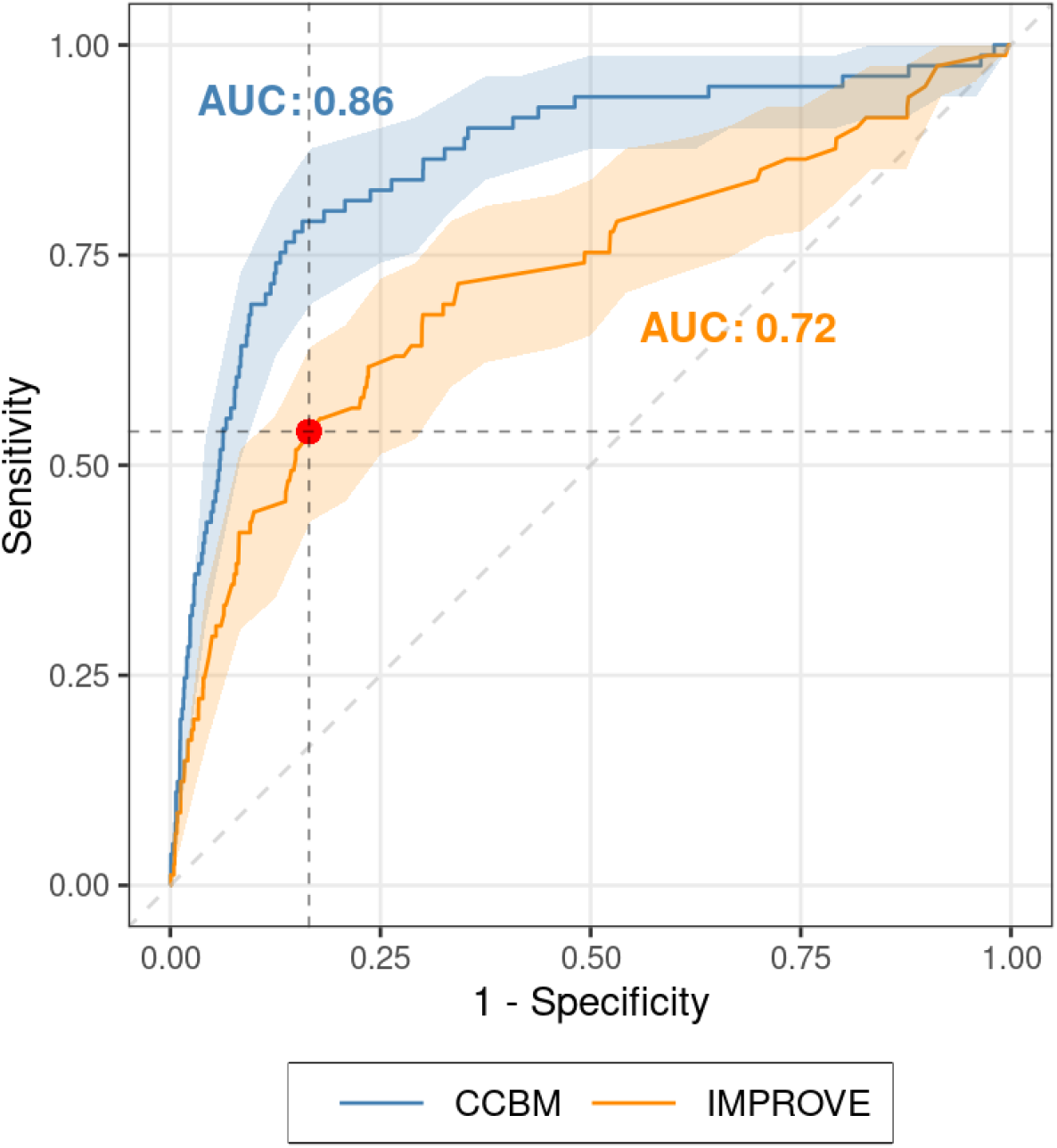
Receiver operating characteristics (ROC) curve comparing the Cleveland Clinic Bleeding Model (CCBM; AUC = 0.86, 95% CI: 0.81-0.91) versus the IMPROVE model (AUC = 0.72, 95% CI: 0.66-0.78) in the validation set (n = 13,895), with bootstrapped 95% confidence bands. The optimal cut-off point for IMPROVE (≥7 points) is indicated by the red dot. Horizontal and vertical dashed lines intersect with CCBM at corresponding operating points given equivalent sensitivity or specificity, respectively.

Next, we matched CCBM and IMPROVE on sensitivity and compared the models on high-vs. low-risk categorization rates. In our validation set, IMPROVE identified 44/81 (54%) of bleeds using the cut-off of ≥7 points. This equated to a risk cutoff of 1.45% using CCBM. At a sensitivity of 54%, IMPROVE categorized 1,676/13,895 (12.1%) of patients as high-risk, whereas CCBM categorized 950/13,895 (6.8%) patients as high-risk. The mean predicted probability of developing a major bleed among patients categorized as high-risk was 4.28% for CCBM and 3.35% for IMPROVE. A chi-square test comparing rates of high-risk vs. low-risk categorization rates between models showed that at a sensitivity of 54%, CCBM categorized significantly fewer patients as high-risk compared with IMPROVE (6.8% vs. 12.1%, χ^2^ = 221.7, p < .001).

We also matched CCBM and IMPROVE on the basis of number categorized as high-risk (12.1%) and compared sensitivity. A chi-square test comparing number of bleeds detected showed that CCBM detected significantly more bleeds than IMPROVE (70% vs. 54%, χ^2^ = 4.44, p = .035).

### Comparing CCBM with Clinician Assessment

A total of 319 patients in the validation set were ineligible for chemoprophylaxis due to clinician assessment of elevated bleeding risk. Among eligible patients, the mean risk for major bleeding by decile, as computed by CCBM, is shown in Figure 3 (left y-axis). The percentage of eligible patients who received VTE prophylaxis in each decile is also shown in Figure 3 (right y-axis). Mean predicted risk increased by decile, with tenth decile patients showing substantially increased predicted risk compared with patients in the first nine deciles. The percentage of patients who received prophylaxis also increased by decile, with the highest risk patients (tenth decile) more likely to have received prophylaxis than the lowest risk patients (first decile).

**Figure 3.**
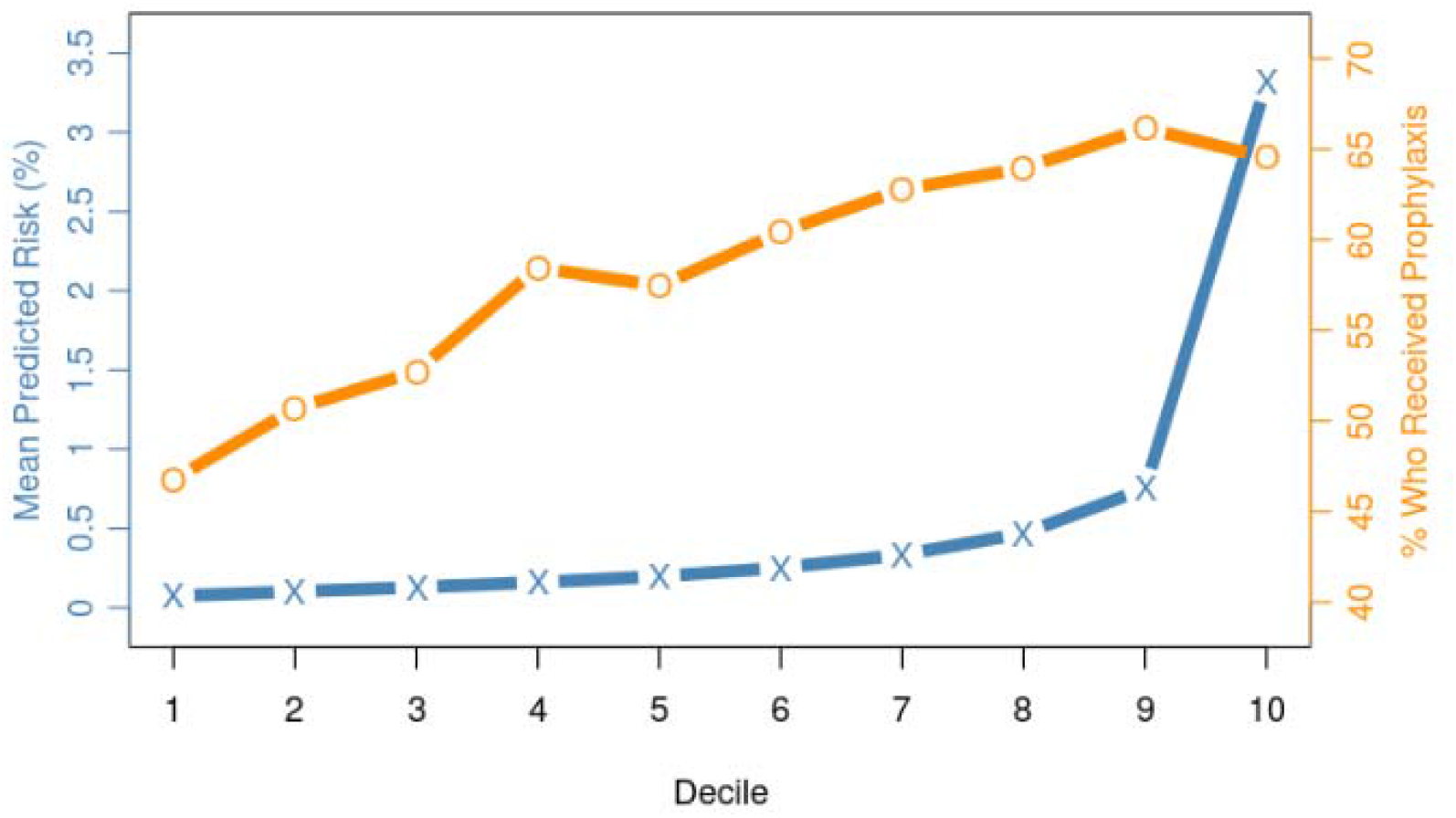
Comparison of mean predicted risk scores computed by CCBM (left y-axis) versus clinician VTE prophylaxis decisions in patients eligible for chemoprophylaxis (right y-axis), by decile of predicted risk.

## Discussion

In this retrospective cohort study, we developed and validated a RAM for major bleeding using data from more than 45,000 patients from 10 hospitals within the Cleveland Clinic Health System. The model, CCBM, contained 17 predictors extracted from the EHR system and demonstrated good calibration and discrimination in the validation set. The CCBM outperformed the IMPROVE^12^ model, which has been the only validated bleeding RAM for more than 10 years. When matching CCBM and IMPROVE on sensitivity, CCBM categorized 44% fewer patients as high risk than IMPROVE, and high risk-patients identified by CCBM had a 28% higher mean major bleeding risk. Thus, if we focus on sensitivity, use of CCBM would prevent as many major bleeds as IMPROVE but allow more patients to receive VTE prophylaxis without increasing their risk of bleeding. Overall, CCBM showed better discrimination than IMPROVE throughout the probable operating range of the ROC curve.

Our RAM differs from IMPROVE in several important ways. First, we had a larger number of patients, which allowed us to identify more risk factors that were independently associated with bleeding and thus improve predictive performance. Second, we restricted the outcome to major bleeds whereas IMPROVE included nonmajor bleeds, which decreases the clinical relevance of predictions because patients at elevated risk of nonmajor bleeding should still receive pharmacological VTE prophylaxis.^27^ Third, we validated both outcomes and all predictors by using both ICD codes and chart review procedures, whereas IMPROVE validated outcomes but relied only on ICD codes to identify risk factors.

Finally, by using a supervised machine learning algorithm, LASSO,^23^ we developed a model that achieved better discrimination in the validation set and is less likely to show degraded performance in future validation sets due to the implementation of variable regularization. By using LASSO, we were also able to identify several additional important risk factors for major bleeding that IMPROVE did not include. Moreover, the IMPROVE model utilizes logistic regression, in which each variable is given the same weight in predicting the outcome. However, LASSO penalizes less important variables, which reduces overfitting and produces less biased predictions in the validation set.^28^ This is important because characteristics vary by region; in our CCHS cohort, peptic ulcers and prior bleeding were the strongest predictors of major bleeding, but these relationships might be weaker in other health systems or patient populations.

Guidelines from both the ACCP and ASH recommend that bleeding risk assessment be performed in conjunction with VTE risk assessment to support VTE prophylaxis decision-making,^4,5^ because patients at increased bleeding risk should not receive pharmacological prophylaxis. But assessing bleeding risk is challenging and clinicians frequently misclassify patients; in our cohort, patients with the greatest risk for major bleeding were more likely to receive prophylaxis than those with the lowest risk. These findings indicate the need for an accurate RAM for major bleeding. The IMPROVE model has been the only validated RAM since 2011, but we have developed a novel RAM that demonstrates better performance. This RAM may be used in conjunction with readily available VTE risk calculators to identify patients at high bleeding risk and decide between mechanical and pharmacological prophylaxis for patients who are at elevated risk of both VTE and bleeding.

Our study makes important contributions but should be evaluated within the context of its limitations. First, although we validated both the outcomes and predictors via chart review, it is possible that we missed some bleeding events or misclassified some of the risk factors due to the imperfect nature of EHR data. Due to the relatively small number of bleeds, it was possible to manually confirm each individual outcome. However, with over 45,000 patients, we could not manually confirm every predictor for each patient. Instead, for each risk factor, we randomly selected a manageable subset of patients and performed iterative chart review until we were confident all variables were as accurate as possible. Second, we had a small number of outcomes to develop and test our model. Our cohort’s major bleeding rate was 0.58%, less than the total bleeding rate in the IMPROVE cohort and external validation studies, which included nonmajor bleeding. Having a highly imbalanced outcome can pose challenges for prediction models and limits the utility of certain evaluation metrics in assessing model performance.^29^

Third, we included patients who received VTE prophylaxis in our cohort. However, our sensitivity analysis showed that most of the variable estimates were not significantly affected by including patients who received prophylaxis. Only peptic ulcer’s effect was meaningfully different, but this risk factor was uncommon in our cohort and thus unlikely to have driven the predictive accuracy of the model. Finally, we did not perform an external validation of our RAM. By splitting our cohort into training and validation sets and adjusting AUC for optimism using bootstrapping, we reduced the bias in our evaluation of model performance. However, the patients in our cohort all came from a single health system, therefore follow-up studies will be needed to validate our RAM in patient populations with different characteristics and distributions of the outcome and risk factors.

## Conclusion

The ACCP and ASH guidelines recommend incorporating bleeding risk assessment into decision-making for VTE prophylaxis, but since 2011 only one bleeding RAM—IMPROVE—has been validated. Using more than 45,000 patient records from 10 hospitals within a single health system, we developed a novel RAM that had good calibration and discrimination in a holdout validation set. Compared with IMPROVE, use of our RAM would allow more patients to receive VTE chemoprophylaxis without increasing their bleeding risk. Future studies should validate the model in external data sets and examine the impact of EHR integration on prophylaxis use, bleeding rates, and other patient outcomes.

## Data Availability

Data produced in the present study are available upon reasonable request to the corresponding author.

## Acknowledgements

This study was supported in part by NIH grants 5T32GM007250-45 and 5TL1TR002549-04.

## Notes

### Competing Interest Statement

The authors have declared no competing interest.

### Funding Statement

This study was funded in part by NIH grants 5T32GM007250-45 and 5TL1TR002549-04.

### Author Declarations

This study was approved by the Cleveland Clinic Institutional Review Board (IRB #14-240).

